# Asbestos as contaminant in the mining of non-asbestos minerals: Case of a “marble” waste slurry dump-yard from Rajasthan, India

**DOI:** 10.1101/2024.10.21.24315894

**Authors:** Raja Singh, Sean Fitzgerald, Rima Dada, Arthur L Frank

## Abstract

Asbestos is fibrous minerals, some of which can naturally occur in certain types of marble, including asbestiform tremolite, actinolite, anthophyllite, and chrysotile, with tremolite the most common among those associated asbestos types. This is of concern as miners and stone workers may not be aware of this and may never be considered by physicians treating them for lung related ailments. Wet marble dust in slurry form is disposed of at designated waste collection points, including in Rajsamand, Rajasthan. unfortunately, this location is now open to the public as a tourist attraction. Dust from this location was tested using Transmission Electron Microscopy (TEM) and tremolite asbestos fibres were found. The presence of tremolite means the workers, bystanders and tourists to the waste collection spots may be exposed to this carcinogen and this may become the etiological factor for mesothelioma or lung cancer among other asbestos related diseases. The marble industry in general should also be subjected to the same regulations as the asbestos industry due to the presence of naturally occurring asbestos in such mines. This is particularly germane in India where asbestos is widely being used and studies show a significant and increasing number of asbestos related lung and pleural, malignant and non-malignant diseases including cancers, such as mesothelioma. This study aims to highlight the occupational and environmental health hazard among workers and the general public.

## Introduction

Regularly, many types of dimension stone are referred to as marble. In reality, marble may be a white coloured stone with the following definition:

*‘a hard crystalline metamorphic form of limestone, typically white with coloured mottlings or streaks, which may be polished and is used in sculpture and architecture’ [1]*

In India marble is widely extracted from the ground and a large part of it comes from the state of Rajasthan. The Geological Survey of India which is run by the Government of India, published a report which stated that total marble recoverable in India is 1200 million tonnes out of which 1100 million tonnes is from Rajasthan [2]. In 1999-2000 4278.63 thousand tonnes of marble was produced in Rajasthan [2]. The location of “marble” deposits of all types in Rajasthan as listed report have been visually represented in Figure 1 below:

**Figure 1.**
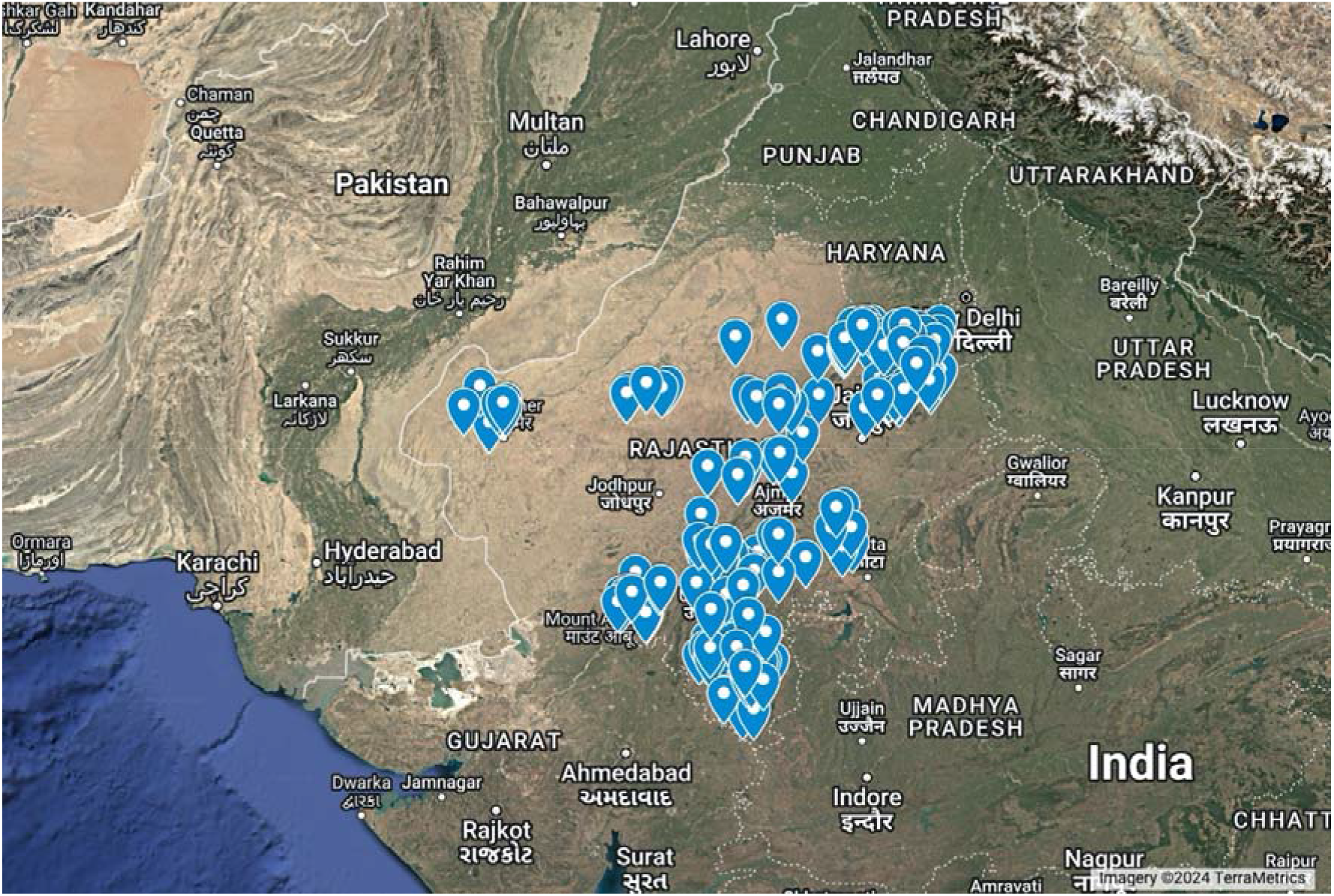
Location of all “marble” deposits which includes pure white marble in the state of Rajasthan in India, with entries with errors excluded. Map prepared by authors on Google My Maps. Information source: Geological Survey of India [2]

India has a standard or marble called IS 1130:1969 [3] which classifies marble according to its colour. The plain white marble is available in districts of Nagaur, Rajsamand, Sirohi, Jaipur, Alwar, Banswara and Udaipur. Marble of this region is Precambrian (older than 540 million years ago), and is significantly metamorphosed; important as the metamorphism from original deposition is conducive to the pressures and temperatures required to produce tremolite as an accessory mineral in the marbles. These marble deposits are part of the Bhilwara, Aravali or Delhi sub-groups. We looked at the Rajsamand District, which has 1637 hectares under marble leases and this is one of the highest in Rajasthan. There are three belts in the Rajasamand District namely Rajnagar-Amet Belt, the Pipali-Kuanria-Dariba Belt and the Kaliwara-Kumbhalgarh-Charbhya Belt. The Rajnagar-Amet Belt consists of the Rajnagar-Bhagwandi-Marchanna block [2]. During the extraction and processing of this marble, there is lot of waste marble that is released in the form of waste marble slurry. It is in the proximity of this block in Rajnagar that there is a marble slurry disposal site, which is described further. Marble waste slurry disposal has been reported to be harmful to the environment despite its economic benefit by another study conducted by the Geological Survey of India in another area, i.e., Makrana (Nagaur) in Rajasthan [4]. The report by geological survey of India from Makrana shows that rills and gulleys were developed over dumped marble wastes. The surface runoff washouts are likely sources for contamination groundwater and surface soil in the areas surrounding the waste dumps. *Calcium, SO4, NO3 and Magnesium was found in soil samples. There was high Total Dissolved salts and SiO2 in the water samples which was derived from mine water pit and the water which was laden with the marble slurry*. [4]

The general assessment of the effects of marble mining in the report shows a positive social economic improvement, but the overall environmental and health cost has a huge potential cost, especially in the long term. Speaking first of the environmental damage, in a report on Makrana marble, by Geological Survey of India, it has been reported that there is severe geoenvironmental degradation of the area. Due to seepage, the ground water was contaminated due to release of waste wate in open spaces, and the water table conditions in one study, due to mining, showed water table dropped below the normal depth and has reached 40-50 metres below ground level. Water in the area also shows high pH, TDS, Ca, Mg, SO_4_, NO_3_, Na, K, Cl, HCO_3_ and SiO_2_. The total impact score was recorded at -3875, which is and impact regarded as *‘Major injurious impact on the environment. Major environmental control measures to be taken and /or site selection for the proposed project to be reconsidered within the buffer zone*.*’ [4]*

In 2023 there was a case before the National Green Tribunal regarding the marble slurry being disposed out of the designated areas in Rajsamand, Rajasthan [5]. On the complaint and on intervention by the local authorities, the slurry was removed, though the method of removal, remediation with dust control and monitoring was not clear. With heavy rainfall, some slurry went outside of designated areas but was removed. But compliance may remain a challenge and may in the future require further complaints by the people affected. Also same slurry was given to local residents for use as household construction material often done by local masons and craftsmen with or without protective equipment or awareness training.

Guidelines for management of marble waste slurry has been developed by the CPCB or the Central Pollution Control Board of the Government of India which have focussed on disposal of marble slurry waste in designated disposal spots. Use of marble slurry as construction material, in cement manufacturing, in synthetic gypsum production, standards in road construction and tile manufacturing has also been promoted by the state pollution board of Rajasthan and also by the CPCB [6,7]. Many studies have been done in this regard [8-13] But currently the solution is the disposal in designated waste collection spots and these waste collection spots may mean large collections at some places, which are open sites which may be against environmental and health norms, but one such site in Rajasthan, i.e. the marble slurry waste deposit spot located in Kelwa in the Mokhampura area or Rajsamand District is not only a simple waste deposit site, but has become a tourist spot in Rajasthan (see figure 2). Rajasthan Tourism promotes this location as a tourist spot with a description: *‘Discover the captivating allure of Rajsamand Dumping Yard! With its vast expanse of white marble slurry, this hidden gem provides a breathtaking backdrop for memorable photoshoots*.*’* [14]. It has also been reported that photographers bring in wedding couples for photo shoots in Rajasthan [15]. The site has an entry ticket and facilities that are being provided by the marble manufacturers association. This is regardless and inconsiderate of the potential negative human health impact as marble slurry is an environmental waste and hazard that contains constituents likely to affect the people who may be exposed to the dust who are being encouraged rather than discouraged to disturb this potentially hazardous dust and debris. This should stop being promoted as a tourist and photographic venue. Interaction with such dust should be kept to a necessary minimum and require training, awareness, and involve use of personal protective equipment and under strict regulations [16].

**Figure 2.**
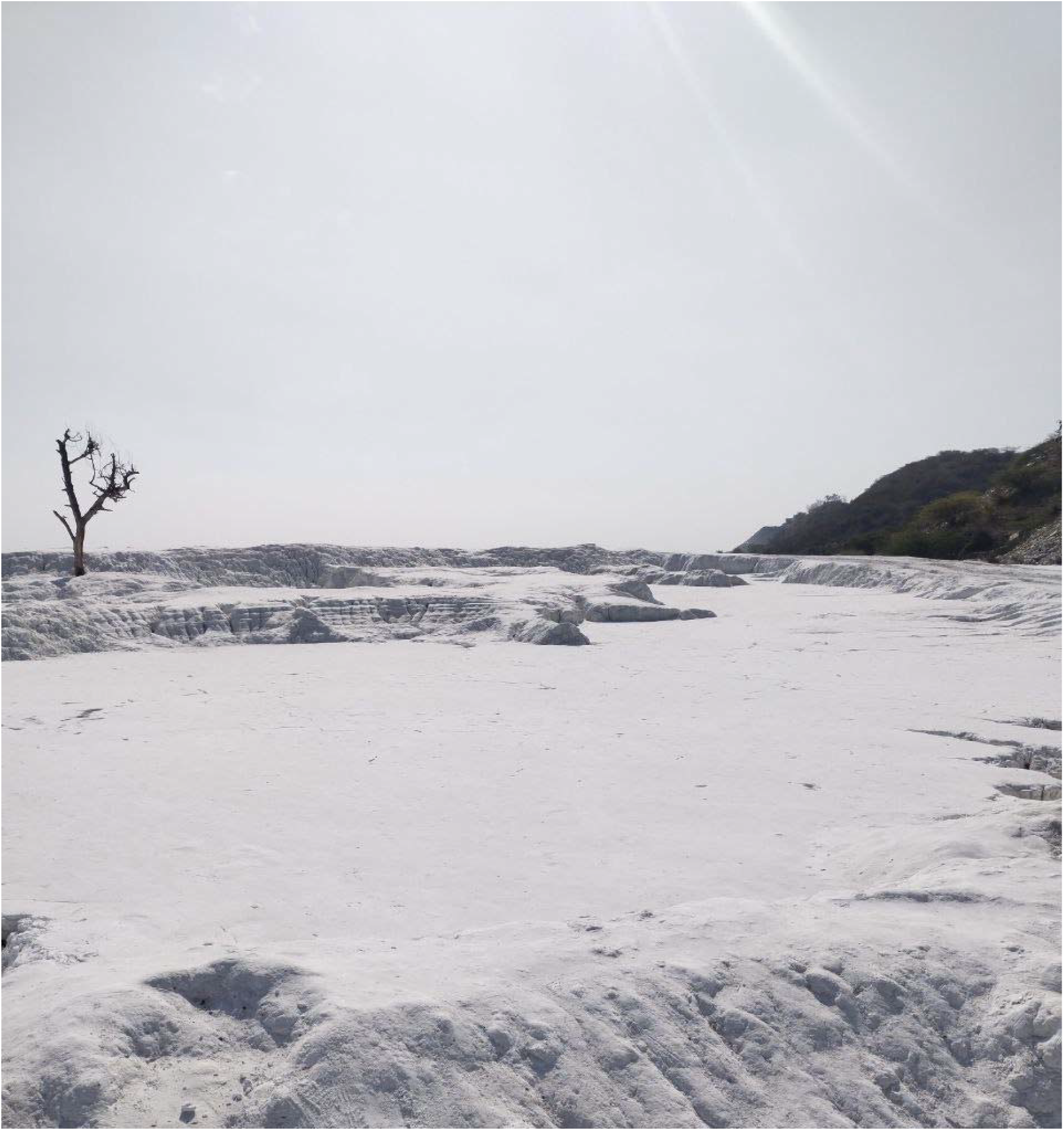
Photograph of the Marble Slurry Waste collection spot near Mokhampura, Kelwa in District Rajsamand, Rajasthan. (Source: Praveen K, Authors)

This marble slurry disposal site is an environmental hazard as it leads to air pollution. In the dry season, the sludge dries up, mixes with wind and deposits on vegetative landscape. The main issue is the impact of marble slurry not only to the environment which has been demonstrated in studies [4], but the potential impact on human health. The slurry when dried, through wind or movement in rains to other areas and subsequently drying, and when used by local residents as construction material, as was evident in the court case above, can cause health effects. It also causes soil pollution and it has been reported by the Geological Survey of India that contractors dispose of slurry in nearby open lands during the night. During rains this slurry spreads and when dry it pollutes the air and travels far beyond the confines of the environment. There is also high air pollution and noise pollution (due to the use of machinery) and the dust deposits on plant leaves and degrades the flora and fauna [4].

These health effects can be substantial when the marble has a natural contamination with asbestos which can include chrysotile, tremolite or actinolite, based on the local geology. In the Rajnagar marble area, where the marble slurry waste disposal in this current study is concerned, the petrographic analysis by the Geological Survey of India states that the marble contains high silica, actinolite, tremolite and diopside [2]. Actinolite and Tremolite are two identified forms of asbestos when fibrous, which are two of the six types of asbestos that are regulated as a known health hazard. Asbestos is the collective term used for 6 types of minerals that have been classified into two categories, amphiboles and serpentine. hrysotile is serpentine asbestos and the amphiboles regulated when fibrous are actinolite, tremolite, crocidolite, amosite and anthophyllite. All varieties of asbestos are carcinogenic [17] and their inhalation can cause asbestosis and malignancies like mesothelioma or lung cancer.

However, petrographic analysis may not provide an indication of a direct health hazard as the ingrained asbestos fibre may not have been released. Hence, it is important to understand whether there is a release of asbestos fibresfrom the marble when it is extracted, cut, finished or transported. In this study, the dust from the marble slurry located near Mokhampur, in Kelwa, District Rajsamand, Rajasthan has been analysed to check for the presence of asbestos.

Aim: The study aims to check asbestos presence in the marble slurry dust that finds its way into the open environment, and to develop appropriate testing protocols and suggest awareness training programs when asbestos from these potential sources is found.

## Methodology

### Materials and Methods

A sample of slurry waste was analysed for characterisation of the constituents, followed by identification of minerals and finally to check for the presence of asbestos by Polar Light Microscopy and Transmission Electron Microscopy as prescribed in the Test Method *EPA/600/R-93/116: Method for the Determination of Asbestos in Bulk Building Material* which is published by the Environmental Protection Agency of the United States[18].

Further, it was assumed that as the product may have the nature that is as the likely to make airborne particles which are respirable-size. Therefore, AHERA or was used to further analyse the dust and to make sure that the structures came under the definition of structures that are countable [19] and if such was found, it could be quantified structures-per-gram weight basis (str/g of asbestos.

Leica DM750P (400x power)petrographic microscope with wave retardation, cross-polarised filters, and dispersion staining techniques at magnifications up to 400X was used for Polar Light Microscopy analysis. Transmission Electron Microscopy analyses were conducted on a JEOL 2000FX (upto 50000x, 100KeV) equipped with Energy dispersive X-ray spectroscopy or EDS and structure by Selected Area Electron Diffraction or SAED.

Preparation for electron microscopy analysis was done by weighing and suspending a portion of the sample in an water and alcohol mix. The suspension was prepped in a manner consistent with *ASTM D6480 Standard Test Method for Wipe Sampling of Surfaces, Indirect Preparation, and Analysis for Asbestos Structure Number Surface Loading by Transmission Electron Microscopy*, i.e., aliquots of measured volume of the sample suspension ollowed by filtration through a 0.2 µm mixed cellulose ester (MCE) filter paper. The final MCE filter was dried, acetone-collapsed and coated with carbon in a vacuum evaporator. The solids and fibres which were collected on the carbon-coated filter replicate were transferred onto copper grids for TEM analysis.

The sample, which is dust sampled from the open ground of the designated waste collection point, was collected in December 2023 and analysed in January of 2024.

The study did not involve any human participants, animal tissue, or animals and hence no ethics review is required for the same. The authors declare the same.

## Results

Analysis of the Kelwa marble dust for mineral content and possible asbestos by PLM did not conclusively detect fibres consistent with asbestos at the light microscope level of detectability. However optical characteristics consistent with the mineral tremolite were observed (*Figure 3*).

**Figure 3.**
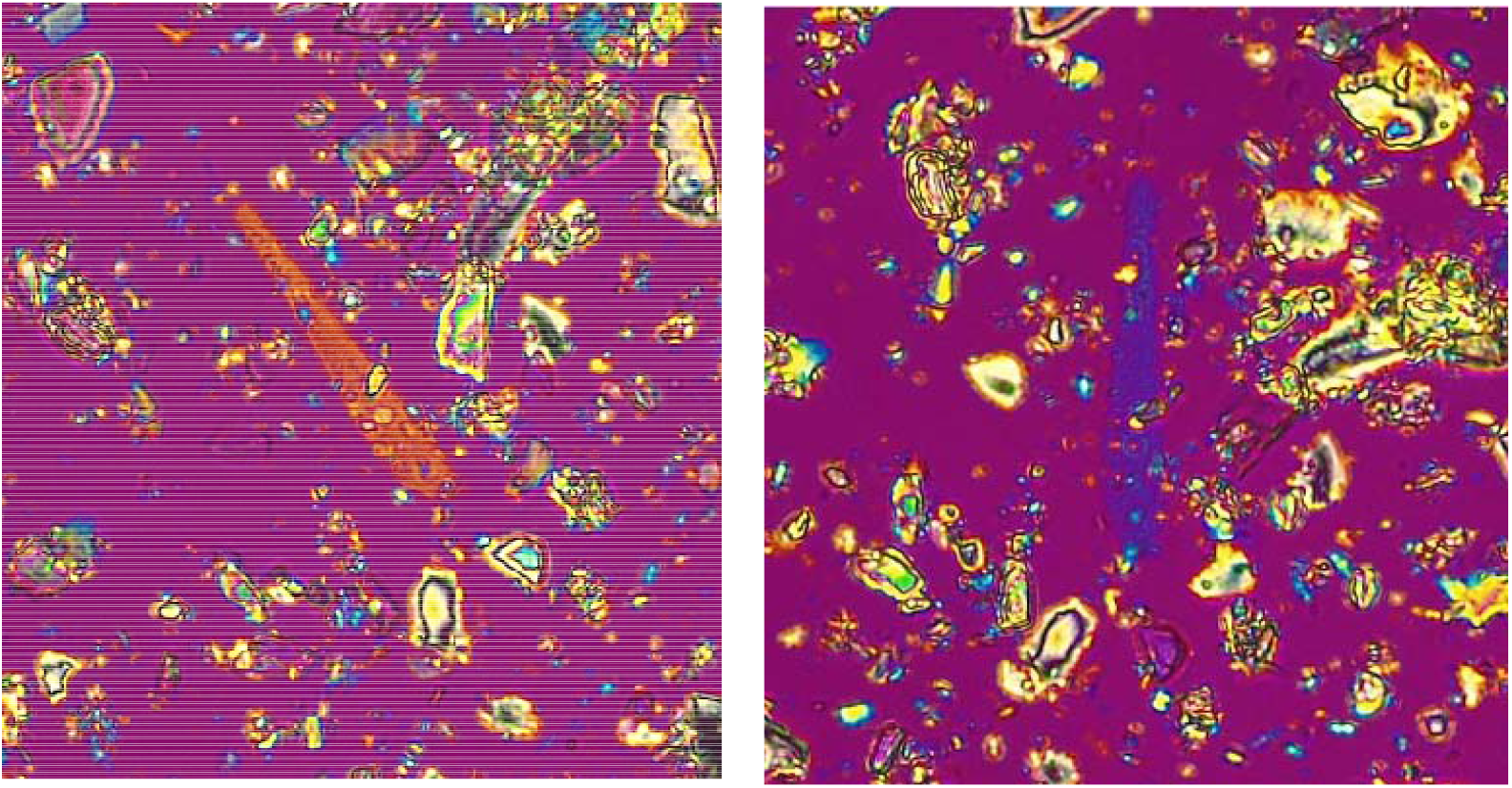
PLM Identification of elongate to fibrous tremolite constituent of Kelwa dust sample. The fibre-bundle rotated to show sign of elongation and oblique extinction characteristics consistent with tremolite, 200x magnification, cross-polarized light with 430nm wave plate compensator. Particles in the Kelwa dust sample also were found consistent with other diagnostic optical characteristics for tremolite, including birefringence and refractive indices, measured in 1.605 HD RI oil particle mounts. Source: Authors.

**Figure 4.**
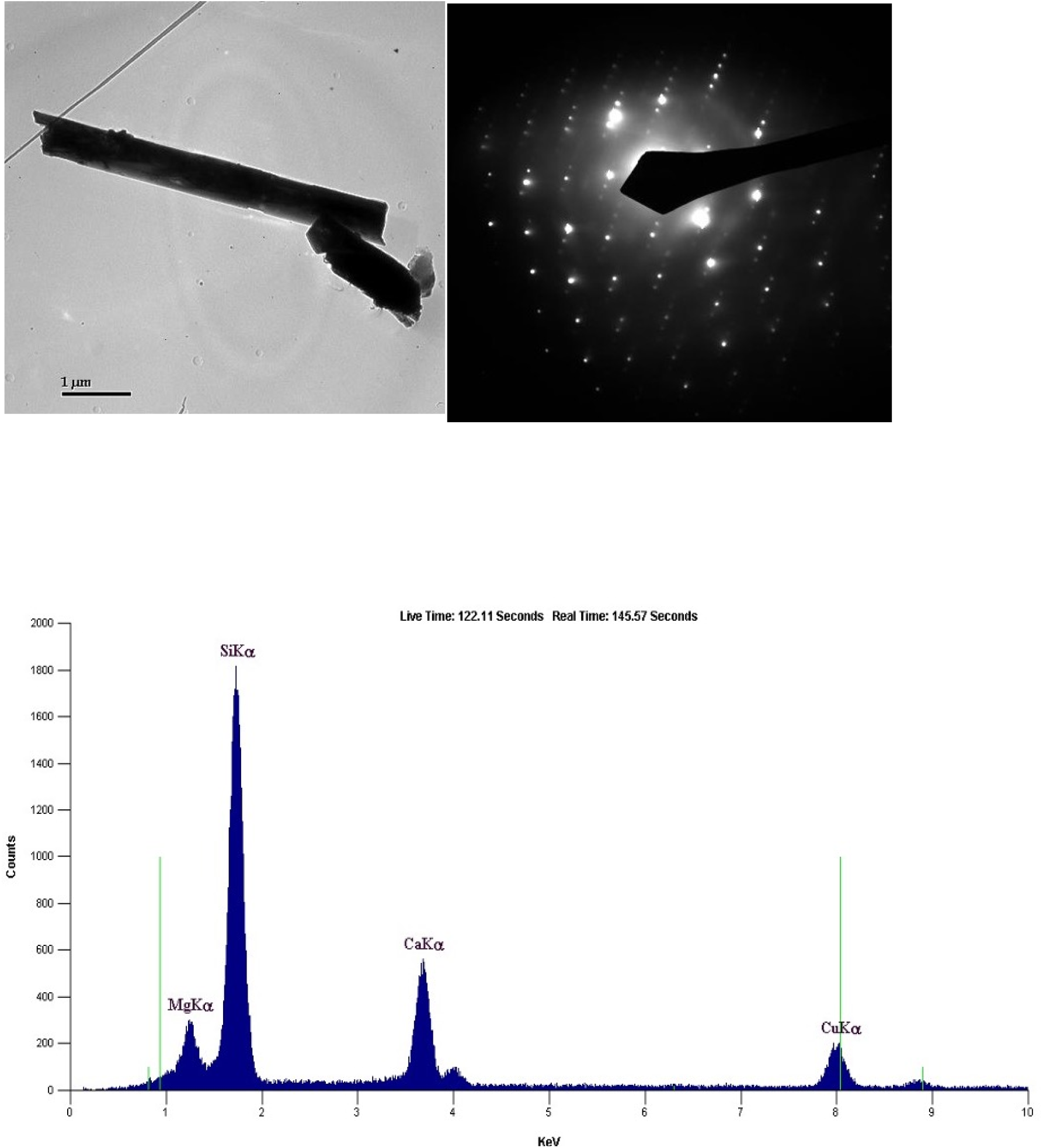
Photomicrograph, diffraction pattern (SAED), and chemistry (EDS) of tremolite structure, µm long containing fibres 0.16 µm wide, bundle width 0.53 µm wide identified in this testing (Marble Dust from Kelwa, Rajasthan, India). Source: Authors

**Figure 5.**
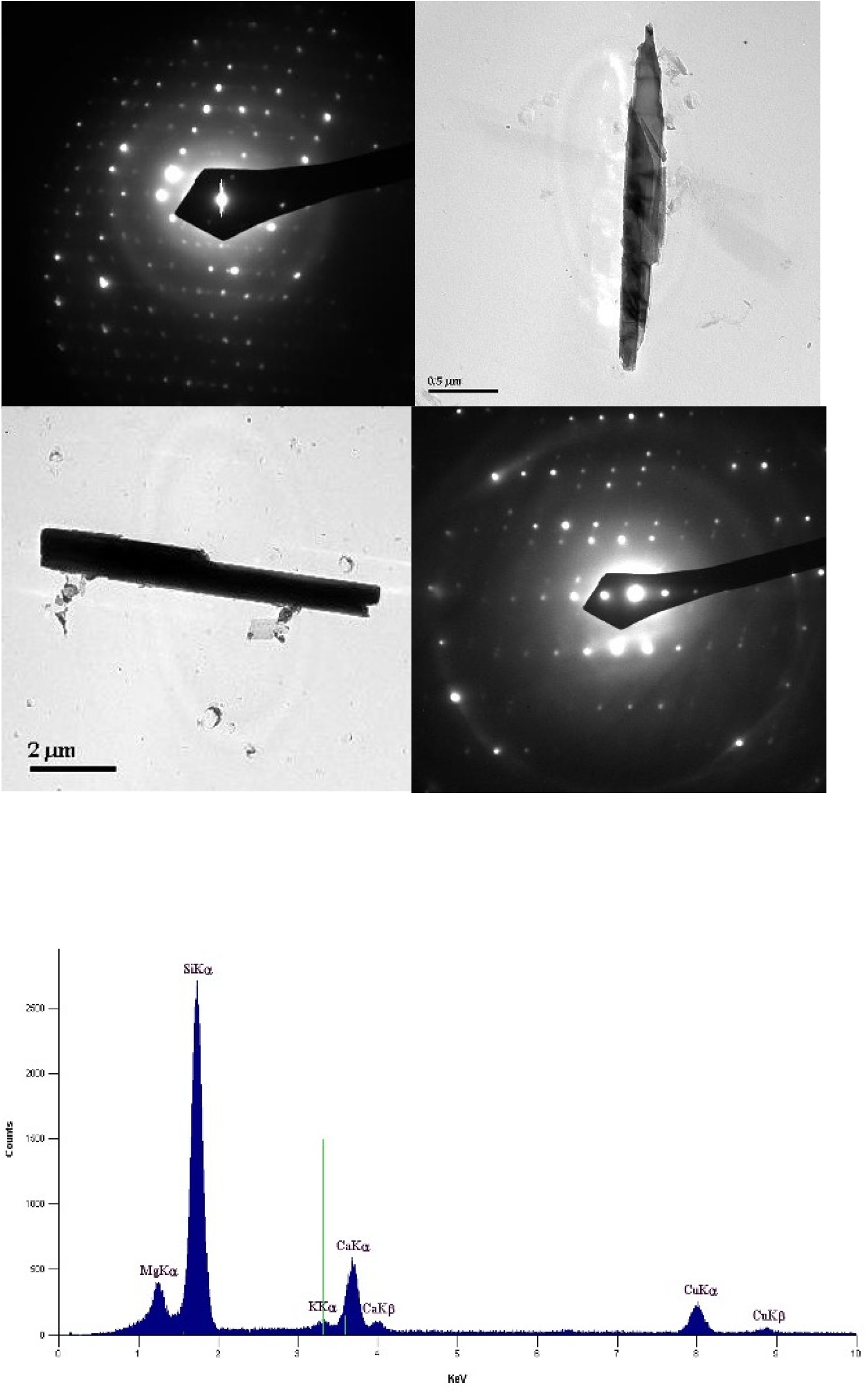
Photomicrographs, diffraction patterns (SAED), and chemistry (EDS) of more countable tremolite structures identified in this testing (Marble Dust from Kelwa, Rajasthan, India). Source: Authors.

**Figure 6.**
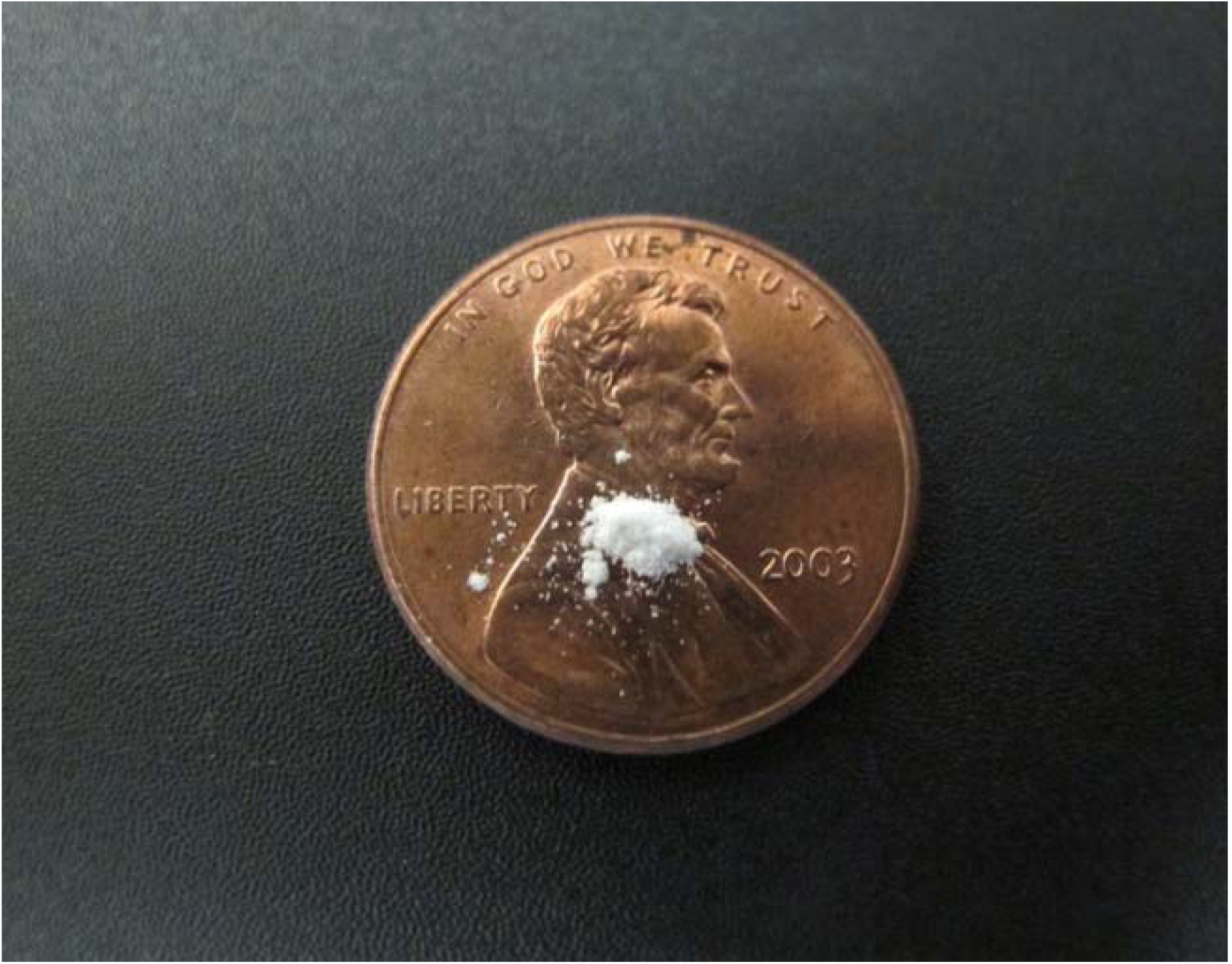
10 milligram of talc for scale on a U.S. penny. An equivalent amount of the Kelwa Marble Dust as tested would contain approximately **37**,**000** countable fibrous structures of the regulated asbestos mineral tremolite. Source: Authors.

Subsequent analysis by TEM confirmed tremolite mineral chemistry by Energy dispersive X-ray spectroscopy or EDS and structure by Selected Area Electron Diffraction or SAED. Tremolite particles were found abundantly dispersed throughout the material, in morphologies ranging from blocky, bladed, and euhedral, with most tremolite particles less than 10 microns, preferentially oriented to elongate, with a substantial constituent identifiable as fibres/fibrous structures which can be counted as asbestos structures. By the AHERA protocol. These would be considered inhalable.

Tremolite asbestos concentration measured in the unit fibre structures per gram (str/g) of the dust was calculated at 11,800,000 str/g for the size fraction >5.0 µm long, and 37,100,000 str/g in consideration of all AHERA-countable asbestos fibre-structures. Adding in all tremolite particle sizes and shapes bumps the tremolite structure count to 103,000,000 str/g.

Form the above, it may be stated that any disturbance of this loose disaggregated dust would more likely than not create a medically significant exposure to asbestos by a person exposed to this in their breathing zone by disturbance of the dust, debris, or soil with this level of asbestiform tremolite concentration.

## Discussion

The initial impetus for this study was a study reported from Udaipur in Rajasthan which documented 76 cases of mesothelioma in 4 years from one hospital [20]. Surprisingly, all of the subjects denied any past exposure to asbestos. However, 69 out of the 76 or 91% reported to have history of direct or indirect exposure to mining and quarrying industry of marble and granite. The study did not investigate for asbestos presence in marble mines but reflected that *‘asbestos is present in close association with marble in rocks and quarrying might lead to airborne asbestos fibres which may be implicated as a causative factor in this region*.*’[20]* This is all the more true as all the subjects were from a mining rich area of Rajasthan where there are marble mines. The current study bridges this information gap by actually testing the marble dust and confirming the presence of tremolite, a type of asbestos which is an etiological factor for mesothelioma. Further, other studies report that protective equipment and precautions are seldom used in mining in Rajasthan, with no or minimal health record-keeping or workers who are in the unorganised sector [21,22].

Studies from other parts of the world were also reviewed. In Namibia, the Kribib area has commercial marble called Rhino White Marble. Amphibolic asbestos, e.g.., tremolite, has been observed in these metamorphic rocks. It is during the extraction or quarrying of the marble that the tremolite can break away and get released into the air.[23]. Importantly, the United Kingdom’s Health and Safety Executive studied marble samples and reported the presence of tremolite fibres in majority of the samples that were studied. But in its document or advisory it stated that this tremolite may be in low quantity of the total volume of the marble and only during cutting or grinding may the asbestos be released[24].

In a study from the US which called for a policy on naturally occurring asbestos, clusters of mesotheliomas were analysed through a literature review in places where direct daily exposure to naturally occurring asbestos was common [25]. This included agricultural villages of New Caledonia, Greece, Turkey, Cyprus and Corsica, where the white soil containing tremolite asbestos is commonly used white stucco which has been widely used there for whitewash the walls or to plater them [26].

There have been other studies from Italy, but which dealt with the presence of chrysotile in Valmalenco serpentinites, but the maximum exposure of asbestos released into the air was near the block cutting which seems to have used multiple blades. The second was at the drilling sites in the quarries[27]. In another study on green “marble” or serpentinite, called Malenco Serpentinite, it was reported that small amounts below 400 ppm were detected in soils and stream sediments along with traces of asbestiform tremolite. Dues to slip fibre mineralisation, a chrysotile layer was expected, but the tremolite was attributed most likely to the talc veins and/or the ophicarbonates that occurred both in serpentinites as well as dolomitic marbles [28].

As to the releasability of asbestos, it may be stated that even low asbestos concentrations may mean potential human health risk [29]. The US Environmental Protection Agency or EPA and the Agency for Toxic Substances and Disease Registry or the ATSDR have studied several sites where low asbestos content produced airborne concentrations of significant concentrations, including sites where asbestos was naturally present. [30,31]. Therefore even fractions a percent of asbestos content by weight leads to proven substantial airborne concentrations which is also the case for the marble “dust” or soil tested here. There has been a debate on this due to the issue of a recent creation of a release index developed in Italy, which creates a process where standard quantity is ground to check the actual releasability, but that particular study used the serpentinite as opposed to the current study uses the actually dust on the ground after release which shows positive asbestos content [32]. Also, mesothelioma was reported from clusters in excess where natural asbestos fibres were found [33].

In Korea, there has been some debate and discussion on the regulation of construction stones which may contain naturally occurring asbestos. Dolomite from a mine in Korea contained asbestos, as well as in the talcum powder for babies available in the Korean market. The Serpentinite from Korea also contains chrysotile. Korea has a ban on asbestos and there is an Asbestos Safety Management Act passed in 2012. But, due to the commercial importance of construction stones, the Korean ministry of labour and employment changed the standard for content of asbestos in materials from 0.1% to 1 percent as it was done to create harmony with the occupational safety law. But it is generally important to consider the naturally occurring asbestos in construction material (apart from materials that have intentionally added asbestos/ industrial products) when regulation of asbestos is concerned as occupational exposure from naturally occurring asbestos is possible, especially when cases of occupational asbestos exposure in Korea increased each year [34].

The lack of regulation, apart from the effect on health and environment, can also mean economic loss to marble miners. Due to regulations in Australia, import of asbestos containing stone ‘knowingly’ even with trace levels is considered a serious offence under the Customs and Workplace Health and Safety (WHS) Laws. It is also reported that Indian Bidasar varieties contain asbestos which may have been imported in Australia from India [35,36]. This means that having proper testing and monitoring of Indian stone is not only important for the sake of occupational health and safety but also to prevent trade prohibitions for Indian materials.

Under-reporting of exposure is a big concern as there can be brief unknowing and para-occupational exposures. A short period of indirect exposure during childhood is often ignored by or is unknown to a leading to under-reporting of the cases or the aetiology. [37]. This may particularly be aggravated when there is a general lack of awareness about the presence of asbestos in a material which may not be gy7y7uysuspected, or such information may not be easily available or circulated. This can be the case of contamination of asbestos in marble which is mined in Rajasthan and elsewhere in India.

Asbestos in India is widely used in the form of building products, where chrysotile fibre is imported and processed in India [38]. There are also major concerns with the non-occupational use of asbestos cement sheets where the end users may be exposed to asbestos fibres due to weathering, demolition, renovation and disposal of asbestos cement roofing sheets [39]. Asbestos has also been reported in the commercial talcum powder in India, which means that millions of people may be exposed to asbestos and this aetiological factor for asbestos related diseases may never occur to physicians[40,41]. There are also ship breaking-cum-recycling industrial unites in India with large number of mesothelioma cases predicted [42]. Apart from the study from Udaipur where 76 cases were reported and possibly linked to asbestos contaminated mining, there have been large studies on mesothelioma in a single hospital in Gujarat [43]. This hospital in Gujarat gets several patients from Rajasthan [44]. Generally with only 16% of the Indian population under the National Cancer Registry Program means that many hospitals are not recording cases in a centralised registry and this is leading to under reporting of mesothelioma cases [44]. This is of much concern as it is predicted that there will be at least 12.5 million patients with asbestos-related in the near future. Further, it is predicted that there will be 1.25 million patients with asbestos-related cancers. In this worldwide cancer figure, half may be from India itself [44]. Under these circumstances, looking at the incidental exposure in mines with asbestos contamination needs urgent review to prevent asbestos related cancers and other diseases in India.

## Conclusion

Asbestos related diseases are significant in India and have been seen to be on a rise across the world. The study aimed to find out whether the waste slurry from a marble waste slurry dump has presence of asbestos. It has been reported in this study where the dust from a marble slurry dumping yard, which is open to the public as a tourist location, has been studied and found to contain tremolite asbestos. This means that the marble industry poses an occupational hazard to the workers, locals around the mine and general public far away, in the form of exposing them to asbestos dust and this may lead to asbestos related diseases which can include the cancer of the lung or the rare malignancy called mesothelioma of the pleura or the peritoneum. This informs us that during extraction process, or during transportation, refining or any other process in and around mines, the tremolite is being released into the environment, into the breathing zone of not only the workers but the bystanders or even tourists. Also, this is all the more crucial as there may not be general awareness about the presence of asbestos contamination in marble which is widely extracted in Rajasthan, and physicians, workers or mine owners may never be aware of the history of asbestos exposure in “marble” mines where is a contaminant.

Recommendations include:

1. There should be wide scale monitoring and testing of the inhalable dust that is released in marble and dimension stone mining in Rajasthan and other areas so that particular lots of the stone can be marked and extra precautions can take place for those lots.
2. Even for general dust precautions worker related protective equipment should be used, but now specially, the personal protective equipment and other dust control methods in mines should be made compulsory and such actions should be monitored and regulated by government agencies. This is valid for marble mines, marble processing areas, marble installation sites and construction sites.
3. Cancer should be made notifiable throughout the country in India as migrant labour may come from states where cancer may not be notified and mesothelioma, a malignancy almost always caused by asbestos can be tracked and reported.
4. There should be a policy in India for naturally occurring asbestos to be identified in all the types of stone where asbestos may be present so that the miners, workers and occupational physicians, pulmonologists, oncologists can include this in history taking and may be aware of the possibility of asbestos related disease when a worker turns up from previously thought non-asbestos mining activity.
5. The marble slurry designated wate deposit spots should stop being promoted as a tourist and photographic venue. Interaction with such dust should be kept to a necessary minimum and require training, awareness, and involve personal protective equipment.
6. When marble is used at homes, buildings, during its laying, the workers and residents may be exposed, during cutting, polishing and grinding, in case it may have naturally occurring asbestos. Awareness is needed and proper guidelines are to be framed for precautions for workers and residents during marble installation. Since marble dust be used for tile manufacture, the same precautions during tile installation may be needed to be in place. This may include use of personal protective equipment like masks and prevention of spread of dust to other areas in the proximity of the construction site. There must be prescription to change work clothes before reaching home for workers to prevent para occupation exposure for partners and family of the workers.
7. Compensation and social security for miners in mines containing naturally occurring asbestos must be treated at par with asbestos industry workers (because mining of asbestos is banned in India [46] and the industry regulations may be relevant) and the directions given by India’s apex judicial body, or the Supreme Court of India must be followed as in the Consumer Education and Research Centre judgment of 1995 [47], by adapting the same for miners. The adapted recommendations for mines having asbestos as contaminant in line with the judgement for asbestos industries may be as follows:
  1. That the health records of the workers in the mines for a minimum period of forty years should be maintained. Or, the health records fifteen years after retirement or end of employment should be maintained, whichever may be later.
  2. Mines with naturally occurring asbestos may undertake membrane filter tests, or its equivalent, to detect fibres of asbestos.
  3. All mine workers should come under Employee State Insurance Act or its equivalent, if not already covered.
  4. The standards for permissible exposure limit to asbestos must be reviewed as per the International Labour Organisation or ILO standards as and when they are released.

## Data Availability

All data produced in the present work are contained in the manuscript

## Acknowledgements

The authors thank the contribution of Praveen K, a geography graduate for the field work in Rajasthan. Special thanks to all the all the people who supported this work directly or indirectly.

## Declarations

Second and last authors do medical-legal work regarding asbestos, primarily for plaintiffs. The other authors declares no conflict of interest. No funding has been received for this work. The first author acknowledges Drexel Dornsife School of Public Health for the post-doctoral fellowship. All data produced in the present work are contained in the manuscript. *This paper has been posted as a prepint on medRxiv server on 22*^*nd*^ *October 2024 [48]*

## CREDiT Statement

Conceptualisation: RS, ALF; Data Curation: RS; Formal Analysis: SF; Investigation: SF, RD; Methodology: RS, ALF; Project Administration: ALF, RS; Validation: SF, ALF, RD; Visualisation: RS, SF; Writing-Original Draft: RS, SF; Writing-review & editing: ALF, RD, SF, RS; Supervision: ALF;

